# Antibacterial consumption in the context of COVID-19 in Pakistan: an analysis of national pharmaceutical sales data for 2019-20

**DOI:** 10.1101/2020.12.05.20244657

**Authors:** Ebiowei Samuel F. Orubu, Farrukh Malik, Albert Figueras, Carly Ching, Ahsan M Fuzail, Veronika J. Wirtz, Muhammad H. Zaman

## Abstract

**Objectives:** Uncertainties in the pharmaceutical management of COVID-19 have led to reports of increased indiscriminate uses of antibiotics. The aim of this study was to investigate this at a national level in Pakistan.

**Methods:** We used IQVIA pharmaceutical sales volume data to analyze antibiotic consumption for January-April 2020 in Pakistan. The consumption of 77 antibiotics for systemic use in 2020 was calculated, and compared against the same period in 2019 to track changes in total consumption and consumption pattern by six assigned therapeutic groups: tetracyclines, quinolones, beta-lactam penicillins, other-beta-lactams, macrolides and “others” – comprising the remaining groups.

**Results:** The total consumption for January-April 2020 was 17.76 Defined Daily Doses per 1000 population per day (a 18.8% decrease from 2019). Quinolones, beta-lactam penicillins, other-beta-lactams, and macrolides constituted 80% of total consumption. There were increases between February-April 2020 of macrolides and “others”. In March 2020, 112768 more people consumed a macrolide per day compared to March 2019; mostly azithromycin.

**Conclusions:** Total consumption decreased in the first four months of 2020 compared to 2019, possibly arising from a combination of several pre-existing and concurrent economic conditions as well as the lockdown situation. However, the results suggest an increase in the consumption of the macrolide class of antibiotics during the early phase of COVID-19 in Pakistan.

## Introduction

The non-evidence based use of antimicrobials during COVID-19 can accelerate antimicrobial resistance (AMR) (1,2). COVID-19 was declared a pandemic by the World Health Organization, WHO, on March 11, 2020, some three months after the December 2019 outbreak in Wuhan. While there is as yet no definite therapeutic intervention specific for the causative agent, Severe Acute Respiratory Syndrome Coronavirus 2, or SARS-CoV2, many medicines have been used off-label, and are currently being evaluated in clinical trials (3,4). In early March 2020, the social media promotion of putative therapeutic antimicrobials led to fears of an increase in community antimicrobial sales and of self-medication which, in some cases, have already led to deaths (5).

Pakistan, the world’s fifth largest country by population, recorded its first COVID-19 case in Karachi, Sindh Province, on February 26, 2020 (6). By April 6, there were only 3277 cases with 50 deaths (6). As of June 25, 2020, these have increased to 192970 cases with 3903 deaths (7). Following the first two cases, one in Sindh province and the other in the neighboring Baluchistan Province in late February, the Baluchistan Regional Government closed schools on February 27, followed by Sindh (8). On March 21, Pakistan imposed a partial national lock-down, suspending schools and public transportation with a stay-at-home order, with movement restricted but for essential supplies and services (9). This was followed by the complete lockdown from April 1 to May 8 (10).

In this study, we test the hypothesis that there was an increase in antibacterial consumption (AbC) during the early stages of COVID-19 in Pakistan. The aim was to analyse AbC during the early spread of COVID-19 in the country from January to April 2020 by comparing against consumption during the same period in 2019, in order to map changes in amount and consumption patterns of antibiotics.

## Methods

### Study design

Using routinely collected pharmaceutical sales data, we analysed AbC in Pakistan according to the well-established Anatomical Therapeutic Chemical/ Defined Daily Dose (ATC/DDD) methodology (11,12).

### Data source

We obtained pharmaceutical sale volume data for January-April of 2019 and 2020 for Pakistan from IQVIA database. This database comprised country-wide distributors pharmaceutical sales volume data in units of product packs sold per month, excluding public hospitals (13). We extracted data for all pharmaceutically used substances, or combinations, of the ATC J01 class (anti-infectives for systemic use) comprising antibacterials only, excluding those for the treatment of tuberculosis. The extracted dataset contained 77 antibiotics available as 4648 products (tablets, capsules, injections/infusions, suspensions/oral powders for suspension and oral drops of different dosage form strengths).

The IQVIA dataset was reclassified into current (2019) ATC codes, and based on a preliminary analysis of sale volumes, recategorized into six therapeutic groups: (I) J01A, tetracyclines, (II) J01C, beta-lactams, penicillins (III) J01D, other beta-lactams, (IV) J01F, macrolides, (V) J01M, quinolones, and (VI) “others”, consisting of eight sub-groups or individual antibiotics not already classified.

### Analysis

Data analysis was performed at the ATC level 3 (therapeutic groups) using two measures of AbC: total consumption and consumption pattern. AbC was analysed as Defined Daily Doses (DDD) per 1000 population per day, or DID (14). For this comparative analysis, we used the estimated population for 2019 of 216565318 and factored in a 2.1% growth to give the population for 2020 as 221113190 (15).

A monthly DID for each antibiotic was calculated from sale volumes using the formula: [Monthly sale amount (mg) x 1000] / [DDD (mg) x days in the month x study population] (16); where monthly sale amount = (Substance dosage form strength (mg/g) x unit pack-size x units sold); and unit pack-size is the number of doses, or standard units, per pack. One standard unit was a tablet, capsule, vial or ampoule. For oral suspensions or syrups meant for use in children, the standard unit was 5 ml (17). For example, the unit pack-size for an oral liquid substance presented as a 60 ml bottle was 60/5, or 12 standard units. With drops, the standard unit was adopted as 1. Thus, a pediatric dropper of 10 ml volume had a unit pack-size of 10. Monthly DIDs were then aggregated by therapeutic group.

Total AbC per year was calculated using the formula above by substituting two terms, namely the ‘monthly sale amount’ with the ‘total monthly sales for all four months’ and the ‘days in the month’ with a time period of 121 for 2020 and 120 for 2019. To analyse temporal changes, we used a “year-on-year” change (%) metric comparing the “yearly” DID for the first four months of 2020 to 2019.

AbC pattern was described in terms of proportions (%) of total AbC by therapeutic group for each month. To analyse differences in consumption pattern we used a month-on-month change (%) metric comparing monthly DID per therapeutic group for corresponding months in both years. All analyses were performed on Microsoft Excel; with validation performed on MATLAB.

## Results

The total AbC for January-April was 17.76 DID in 2019 and 15.08 DID in 2020. AbC was dominated by four groups which comprised about 80% of total consumption: J01C, beta-lactams, penicillins; J01D, other beta-lactams; J01F, macrolides; and J01M, quinolones. Overall, the quinolones were most commonly consumed, forming 23-27% of total consumption.

The “year-on-year” change was -18.8%, reflecting a drop in consumption of J01 antibiotics for the first four months of 2020, compared to 2019. Similarly, there was a general drop in month-on-month consumption across therapeutic groups, except for macrolides and “others” (Table 1).

**Table 1.** Antibiotic consumption for January-April of 2019 and 2020 in Pakistan expressed as defined daily doses per 1000 inhabitants per day (DID) and month-on-month changes in consumption (see Methods).

| Therapeutic group |  | Antimicrobial consumption<br>(DDD per 1000 population<br>per day, or DID) |  | Changes in<br>consumption<br>(%) |
| --- | --- | --- | --- | --- |
|  |  | 2019 | 2020 | Month-on-month |
| **JO1A Tetracyclines** | Jan | 2.31 | 1.96 | -15.14 |
|  | Feb | 2.35 | 1.90 | -19.29 |
|  | Mar | 2.02 | 1.44 | -28.71 |
|  | April | 2.12 | 1.50 | -29.17 |
| **JO1C Beta-lactams, penicillins** | Jan | 4.83 | 3.36 | -30.30 |
|  | Feb | 4.79 | 3.63 | -24.31 |
|  | Mar | 3.93 | 3.70 | -5.75 |
|  | April | 3.53 | 3.18 | -9.92 |
| **JO1D, Beta-lactams, others** | Jan | 3.56 | 3.29 | -7.51 |
|  | Feb | 3.60 | 3.52 | -2.29 |
|  | Mar | 3.38 | 3.00 | -11.30 |
|  | April | 3.47 | 2.36 | -31.89 |
| **J01F Macrolides** | Jan | 3.32 | 3.24 | -2.36 |
|  | Feb | 3.28 | 3.38 | 3.26 |
|  | Mar | 2.91 | 3.42 | 17.58 |
|  | April | 2.67 | 2.48 | -7.14 |
| **JO1M Quinolones** | Jan | 4.75 | 4.16 | -12.42 |
|  | Feb | 5.05 | 4.41 | -12.67 |
|  | Mar | 4.52 | 3.93 | -13.00 |
|  | April | 4.77 | 3.41 | -28.48 |
| **Others** | Jan | 2.15 | 0.95 | -55.62 |
|  | Feb | 1.70 | 1.03 | -39.23 |
|  | Mar | 1.12 | 1.00 | -10.47 |
|  | April | 0.94 | 1.06 | 12.62 |

In February and March of 2020, macrolides consumption rose by 3 and 18% respectively, while “others” rose by 13% in April 2020 compared to the same periods in 2019. For macrolides, the higher consumption, in March 2020, of 3.42 DID from 2.91 in March 2019, meant approximately 112768 additional people consumed the macrolide class per day in the month of March in 2020 compared to 2019; while “others” rose to 1.06 DID in April 2020 from 0.94 DID in 2019 (Figure 1).

**Figure 1.**
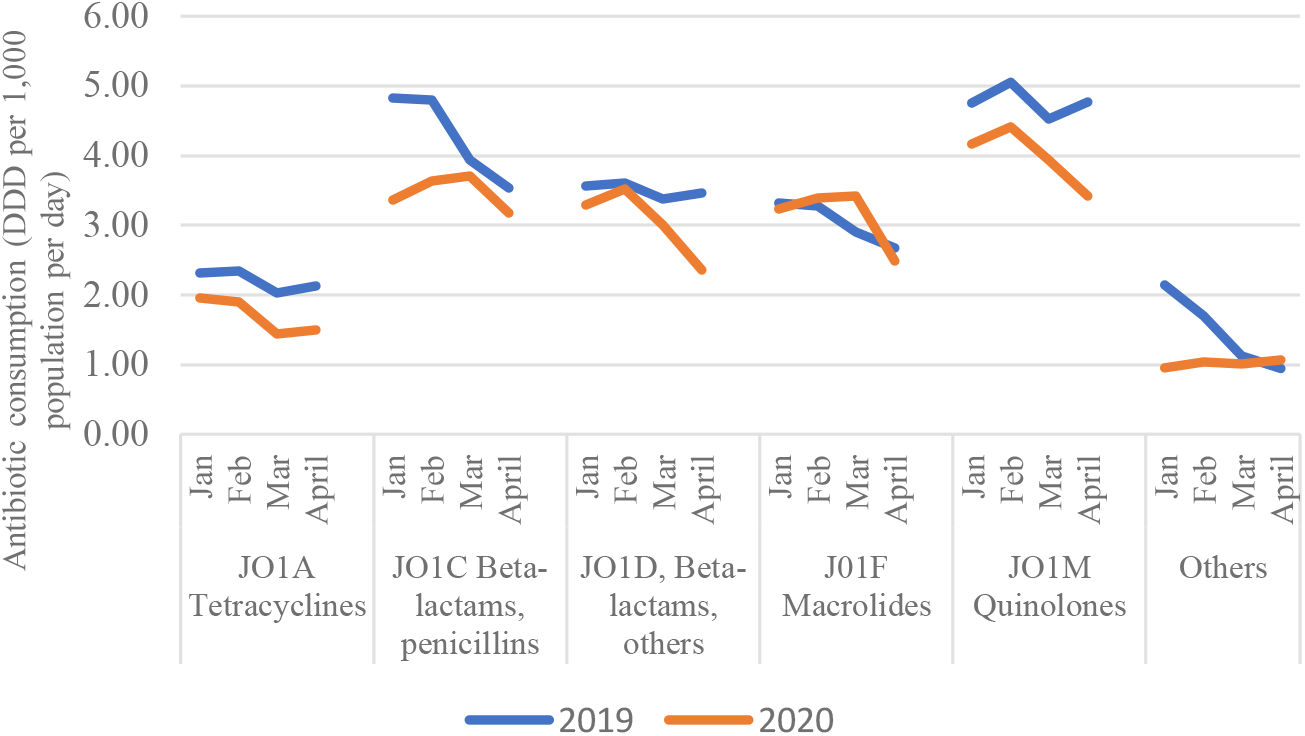
Antibiotic consumption patterns for January-April of 2019 and 2020 in Pakistan. 2020 consumption was lower for antibiotics for systemic use (ATC J01) during the study period of January-April, except for macrolides which showed a rise for February and March of 2020, and “others” with an increase in April 2020.

Within the macrolides – azithromycin, clarithromycin, clindamycin, erythromycin, lincomycin and roxithromycin – azithromycin was the most commonly-consumed. In March 2020, azithromycin contributed 50% to macrolide sale volume with a related consumption of 1.70 DID, compared to 42% of sales and 1.20 DID, in March 2019. Within “others” the rise was driven by increase in the consumption of sulfamethoxazole-trimethoprim.

## Discussion

One of the characteristics of the present outbreak of the COVID-19 pandemic is the general medical uncertainty due to the fact that both the agent (SARS-CoV2) and the disease were completely unknown six months before this study was conducted. Within this context of uncertainty, lack of sound evidence and an unprecedented amount of circulating information, to analyse the consumption of a few antibiotics involved in this information overload is of interest because of the impact of inappropriate consumption on the global AMR threat.

This analysis revealed a “year-on-year” drop in total AbC in the first four months of 2020. This may have due to an economic crisis with currency devaluation and price increases that started in 2018 and supply chain disruptions from these and border closures with China and India in 2019, coupled with COVID-19 related decrease in outpatient attendance in hospitals (18,19).

In contrast, the rise in the consumption of macrolides in March, and, in April 2020, of “others” antibiotics, may have been a result of stock-piling by medicine outlets or clients of azithromycin, following its social-media promotion, alongside chloroquine and hydroxychloroquine, as a preventive or treatment for COVID-19. While “others” also rose, this increase was smaller, being about three orders of magnitude lower.

Within the context of COVID-19, three classes of antibacterials are of interest: macrolides, quinolones, and aminoglycosides. In addition to social media promotion, several early guidelines supported the use of these classes based on studies (20–22). Interestingly, macrolides and quinolones are categorized as “Watch” antibiotics according to the WHO AWaRe classification(23). “Watch” antibiotics are first- or second-choice antibiotics indicated for specific, limited number of infections, with the greater potential to induce resistance even with a single use, and, therefore, prioritized as targets of stewardship programs and monitoring. Thus, the observed high levels of use of fluoroquinolones is troubling. Of all Watch therapeutic groups, the fluoroquinolones, which are the most potent inducers of resistance, was the most commonly consumed antimicrobial class during the study period in Pakistan. Another concern raised by the consumption data analysed in this study is that it represented possible use by outpatients of potential treatments (including azithromycin) for secondary bacterial infections in COVID-19 that should be reserved for hospitalized patients.

## Conclusion

This analysis provides relevant evidence for the national antibiotic stewardship program. Overall, the total consumption decreased in the first four months of 2020 compared to the same study period in 2019, possibly arising from a combination of several pre-existing and concurrent economic and health conditions. However, the results suggest an increase in the consumption of the macrolides during the early phase of COVID-19 alongside a high consumption of quinolones, raising concerns about an increased risk of resistance.

## Data Availability

Relevant data for this work is included. Proprietary data belonging to IQVIA is referenced in the text.

## Acknowledgement

We thank PharmEvo (Pvt.) Ltd. for supporting this study with IQVIA data. This work was made possible by support from the Boston University Social Innovation on Drug Resistance (SIDR) Postdoctoral Program to ESFO and the U.S. Pharmacopeial Convention (USP) Quality Institute to CC.

